# Disentangling fine-scale effects of environment on malaria detection and infection to design risk-based disease surveillance systems in changing landscapes

**DOI:** 10.1101/2020.04.15.20065656

**Authors:** Kimberly M Fornace, Ralph A Reyes, Maria Lourdes M Macalinao, Alison Paolo N Bareng, Jennifer S Luchavez, Julius Clemence R Hafalla, Fe Esperanza J Espino, Chris J Drakeley

## Abstract

Landscape changes have complex effects on malaria transmission, disrupting social and ecological systems determining the spatial distribution of risk. Within Southeast Asia, forested landscapes are associated with both increased malaria transmission and reduced healthcare access. Here, we adapt an ecological modelling framework to identify how local environmental factors influence the spatial distributions of malaria infections, diagnostic sensitivity and detection probabilities in the Philippines. Using convenience sampling of health facility attendees and Bayesian latent process models, we demonstrate how risk-based surveillance incorporating forest data increases the probability of detecting malaria foci over three-fold and enables estimation of underlying distributions of malaria infections. We show the sensitivity of routine diagnostics varies spatially, with the decreased sensitivity in closed canopy forest areas limiting the utility of passive reporting to identify spatial patterns of transmission. By adjusting for diagnostic sensitivity and targeting spatial coverage of health systems, we develop a model approach for how to use landscape data within disease surveillance systems. Together, this illustrates the essential role of environmental data in designing risk-based surveillance to provide an operationally feasible and cost-effective method to characterise malaria transmission while accounting for imperfect detection.

## Background

Malaria transmission is highly variable spatially, driven by the geographical distribution of human populations, mosquito vectors and the environments in which these populations interact (1). Surveillance systems aim to identify these high-risk locations and populations in order to effectively plan, implement and evaluate control policies and measures. As control programmes reduce incidence and transmission decreases, spatial heterogeneity becomes more pronounced, necessitating increasingly higher resolution data to detect foci of infection (2). This becomes increasingly critical in rapidly changing environments, where changing human populations and vector habitats may cause significant shifts in transmission patterns (3). However, despite extensive research linking landscape with malaria transmission, landcover data rarely informs malaria surveillance systems.

For passive surveillance systems relying on reported malaria case data, understanding spatial distributions of risk is further challenged by underreporting due to health-seeking behaviour or asymptomatic infections present in the community. Increasing evidence suggests the proportion of asymptomatic malaria infections not detectable by standard diagnostics increases in low transmission settings, resulting in large numbers of infections not detected by passive surveillance systems (4, 5). These asymptomatic infections are commonly seen in older age groups, with potentially different risk factors and spatial distributions from clinical malaria cases (6, 7). Population-based community surveys remain the gold standard for assessing spatial patterns of infection; however, these sampling approaches are highly resource intensive and may require very large sample sizes in low transmission settings. Alternatively, more operationally feasible surveys of easy access groups, such as health facility attendees or school children, are used to increase probability of detecting infections within the community (e.g. (8-11)). However, both approaches targeting clinical cases and easy access groups are inherently biased and do not fully capture the spatial distribution of infections.

Within ecology, estimates of species distribution or abundance are similarly challenged by imperfect detection and spatially biased observation processes (12). Occupancy models are widely used to estimate the probability that a species occupies a geographic location within a specified time period while accounting for possible non-detection (13). This method partitions observation processes determining detection probability and biological processes determining occupancy probability, each associated with potentially overlapping spatial and environmental covariates. This makes the simple assumption that a species cannot be detected if it is not present; however, if present, the species may or may not be detected during sampling. In addition to allowing estimation of true occupancy states as a latent variable, this method allows quantification of uncertainty in the observation process under different sampling scenarios (14-16).

Here, we combine occupancy models with practical health facility surveys and molecular diagnostics to evaluate the spatial coverage of surveillance approaches and their ability to detect locations with recent malaria transmission. By estimating a spatially explicit sensitivity of different diagnostic methods, we illustrate how environmental data can be used to develop operationally feasible risk-based surveillance systems to increase the probability of detecting areas of infection while rationalising scarce resources. This provides an adaptable framework to integrate convenience sampling approaches into existing disease surveillance systems to target control measures and characterise spatial and environmental drivers of infection from opportunistically collected data.

Using rolling cross-sectional health facility-based surveys in which all attendees regardless of patient status or symptoms were screened for malaria using routine and molecular diagnostics, with residences geolocated in real-time using tablet-based applications, we apply this approach to describe the spatial distribution of malaria infections in Rizal, Palawan, The Philippines (Figure 1 (11)). While the Philippines has made substantial progress towards malaria elimination, with 50 provinces declared malaria free, Palawan contributes over 95% of malaria cases nationally, including 2718 cases from the municipality of Rizal in 2018 (17). Deforestation rates have increased markedly within Rizal; of the 24% decrease in forest cover between 2000 - 2018, over 50% of deforestation occurred after 2015, largely driven by agricultural expansion (18). Within this region, malaria risks are strongly associated with proximity to forested areas due to both vector ecology and occupational risks (19). Described as "frontier malaria,” factors associated with increased malaria risks, such as proximity to forest edges and recent deforestation, are also associated with reduced healthcare access, resulting in potentially complex interactions between detection bias and infection risks (20).

**Figure 1.**
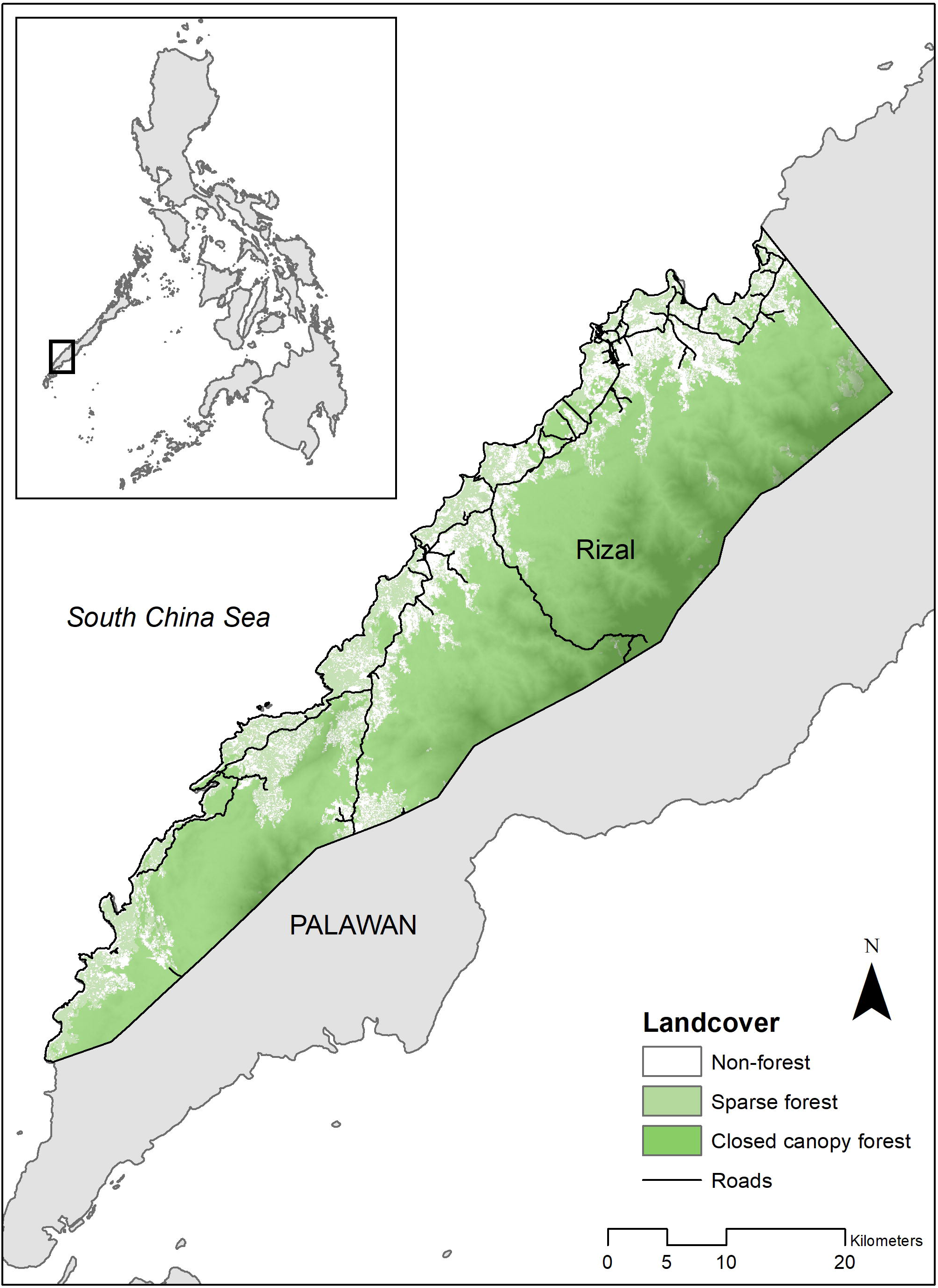
Study area and forest cover in 2017

Our results illustrate that enhanced health facility-based surveys increase the probability of detecting locations with malaria infections by markedly increasing the spatial coverage of the surveillance system in addition to simply increasing the numbers of individuals screened. By comparing locations of infections detected by routine diagnostics (microscopy and/ or rapid diagnostic tests (RDTs)) with sub-patent infections only detectable by molecular methods, we show the sensitivity of routine diagnostics decreases in highly forested areas, with many locations of malaria transmission only detectable by molecular methods. We demonstrate how these findings can be used to develop operationally feasible and cost-effective environmentally targeted risk-based surveillance methods and prioritise locations with high probabilities of infection not captured by existing surveillance systems.

## Results

### Impact of survey method on detection probability

We conducted monthly rolling cross-sectional surveys at 27 health facilities across Rizal, Palawan over a one-year period (Figure S1). All consenting individuals, regardless of symptoms or patient status, were screened for malaria using microscopy or RDTs and polymerase chain reaction (PCR) as described by (21). As multiple malaria species are present in this area and over 75% of infections were with *Plasmodium falciparum*, we classified malaria as positive for any *Plasmodium* species. Using this data, we initially compared two surveillance approaches: 1. Standard - passive case detection (PCD), including febrile patients screened using routine diagnostics (RDTs or microscopy) as per current national surveillance guidelines; 2. Enhanced - standard PCD plus screening all health facility attendees with both routine and molecular methods.

Between June 2016 - June 2017, 5767 individuals were enrolled in this study, including 1914 (33.2%) febrile patients screened for malaria by existing passive surveillance systems (21). Of these, 801 individuals (13.9%) were positive for malaria by molecular methods and 498 were positive by RDT or microscopy. We geolocated all residences in Rizal (n=7313), identifying individuals screened by PCD from 698 unique locations while enhanced surveillance screened individuals from 2201 locations (Table S2). Malaria infections were detected at 352 household locations using enhanced surveillance and 117 locations by standard PCD.

As control measures are targeted based on reported locations with malaria infection and over 80% of infected locations had only one infected individual detected, we chose to model whether malaria infection was present or absent in a specific location (occupancy) rather than incidence. Detection probabilities, the probability of screening at least one individual from a location during the study period, varied geographically, with travel time to the nearest health facility negatively associated with detection probability by both PCD and enhanced surveillance methods (Table 1). Enhanced surveillance increased detection probabilities over three-fold compared to standard PCD (mean probability 3.34, 95%BCI: 1.03 - 8.27) in addition to markedly increasing spatial coverage of surveillance, particularly in rural populations living in forested areas (Figure 2A, 2B).

**Table 1.** Posterior estimates of fixed effects and spatial range for joint models of A. standard PCD and B. enhanced surveillance

|  | Mean | SD | 95% BCI |
| --- | --- | --- | --- |
| **Probability of detection** |  |  |  |
| Distance to roads | 0.226 | 0.125 | -0.020, 0.227 |
| Travel time to clinic | -0.317 | 0.120 | -0.561, -0.090 |
| Distance to forest | -0.112 | 0.053 | -0.217, -0.009 |
| Spatial range (km) | 8.140 | 2.500 | 4.319, 14.050 |
| **Probability of infection** |  |  |  |
| Distance from roads | 0.094 | 0.101 | -0.109, 0.287 |
| Population density | -0.603 | 0.145 | -0.894, -0.322 |
| Precipitation of wetness month | 0.212 | 0.107 | -0.006, 0.420 |
| Distance from closed canopy forest | -0.222 | 0.156 | -0.537, 0.078 |
| Spatial range (km) | 1.752 | 1.096 | 0.493, 4.643 |
| Scaling parameter for shared spatial effect | 0.590 | 0.163 | 0.282, 0.921 |

\*\* All covariates mean-centred and scaled

B.
|  | Mean | SD | 95% BCI |
| --- | --- | --- | --- |
| **Probability of detection** |  |  |  |
| Population density | -0.533 | 0.085 | -0.701, -0.368 |
| Travel time to clinic | -0.511 | 0.107 | -0.729, -0.309 |
| Aspect | 0.094 | 0.039 | 0.017, 0.171 |
| Spatial range (km) | 15.804 | 6.130 | 7.212, 30.957 |
| **Probability of infection** |  |  |  |
| Distance from roads | 0.285 | 0.071 | 0.145, 0.423 |
| Upslope area | 0.181 | 0.111 | -0.038, 0.399 |
| Topographic wetness index | -0.243 | 0.120 | -0.481, -0.011 |
| Temperature annual range | 0.236 | 0.098 | 0.043, 0.428 |
| Distance from closed canopy forest | -0.326 | 0.111 | -0.548, -0.112 |
| Spatial range (km) | 0.897 | 0.232 | 0.532, 1.438 |
| Scaling parameter for shared spatial effect | 1.216 | 0.177 | 0.887, 1.179 |

\*\* All covariates mean-centred and scaled

**Figure 2.**
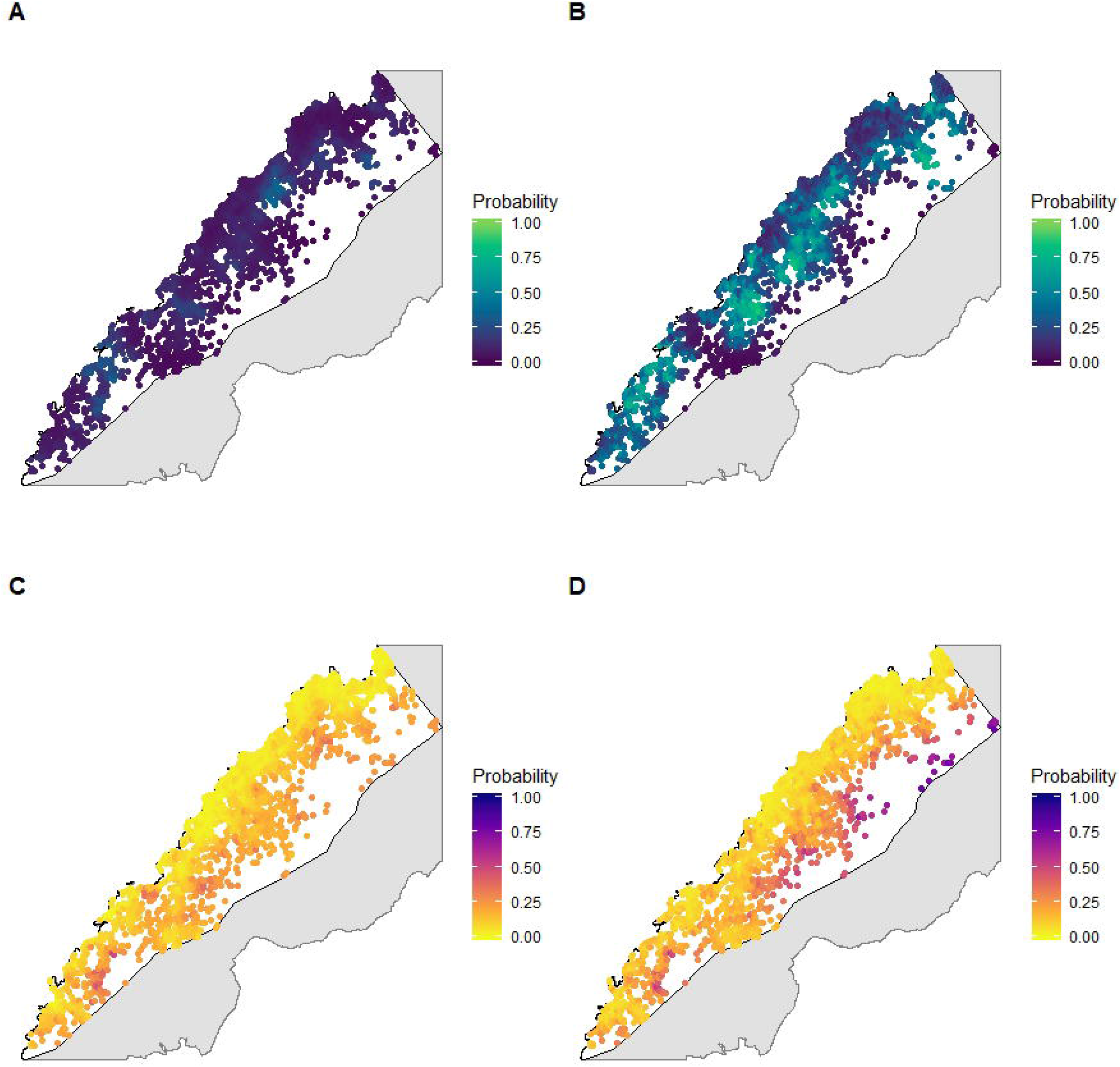
Posterior probability of infection under different sampling scenarios adjusted for detection probabilities: A. detection probability using routine passive case detection; B. detection probability using health facility-based surveys; C. probability of infection estimated from passive case detection using routine diagnostics; D. probability of infection estimated from active case detection using molecular diagnostics

### Spatial distribution of infection

Incorporating these detection probabilities into hierarchical occupancy models revealed a much wider spatial distribution of malaria detected by enhanced surveillance compared to PCD alone, identifying areas of infection not captured by existing surveillance systems (Figure 2C, 2D). We identified a range of differing spatial and environmental risk factors for infections detected by different diagnostic methods; however, all infections were associated with proximity to closed canopy forests in more rural populations (Table 1). For joint models for each surveillance scenario, incorporating a shared spatial random effect between detection and infection probability improved model performance, suggesting a common spatial process driving healthcare access and disease risks (Table S3).

To explore the factors determining these differing distributions of infections, we estimated the probability of patent malaria detectable by RDT or microscopy in all malaria infections we identified. Malaria infected individuals were identified from 435 locations and over one third of infected individuals (37.8%, 95% CI: 34.5-41.3%) could only be detected by molecular methods. Subpatent malaria was substantially more common in forested areas with the odds of patent infections increasing 1.23 (95%BCI 1.03-1.47) with every kilometre distant from closed canopy forested areas (Table 2). Using data from all residence locations identified within Rizal, we predicted a location-specific probability of patent malaria, equivalent to the sensitivity of routine diagnostics (RDT and/or microscopy) (Figure S2).

**Table 2.** Posterior estimates of fixed effects for malaria positive households detected by routine diagnostics (RDT and microscopy) compared to molecular methods

|  | Mean | SD | 95% BCI |
| --- | --- | --- | --- |
| Distance from closed canopy forest* | 0.225 | 0.112 | 0.036, 0.476 |
| Annual precipitation* | -0.318 | 0.121 | -0.620, -0.145 |
| Precipitation of wettest month* | 0.256 | 0.085 | 0.091, 0.422 |
| Population density<sup>2*</sup> | 0.093 | 0.087 | -0.078, 0.263 |

\* Mean-centred and scaled

### Evaluating surveillance systems

As our primary aim was to identify all locations of malaria infection, we evaluated surveillance approaches against estimates of true infection status using molecular diagnostics and all available spatial data. Accounting for spatial bias of collection data, we estimated 11.4% (95%BCI: 4.6%-21.9%) of all locations in Rizal had malaria infections during the sampling period. While no surveillance method identified all areas of infection perfectly, enhanced surveillance using molecular methods identified 38.7% (95% BCI: 33.6-43.8%) of infected locations while PCD only identified 5.7% (95% BCI: 0.1 - 11.9%). Including molecular diagnostics in PCD only slightly improved the probability of detecting infections while conducting health facilities using only routine diagnostics increased the number of infected locations identified slightly more (Figure 3A, Figure S4). We additionally identified 247 locations with a very low (<0.05% probability) of detection by any health facility-based surveillance.

**Figure 3.**
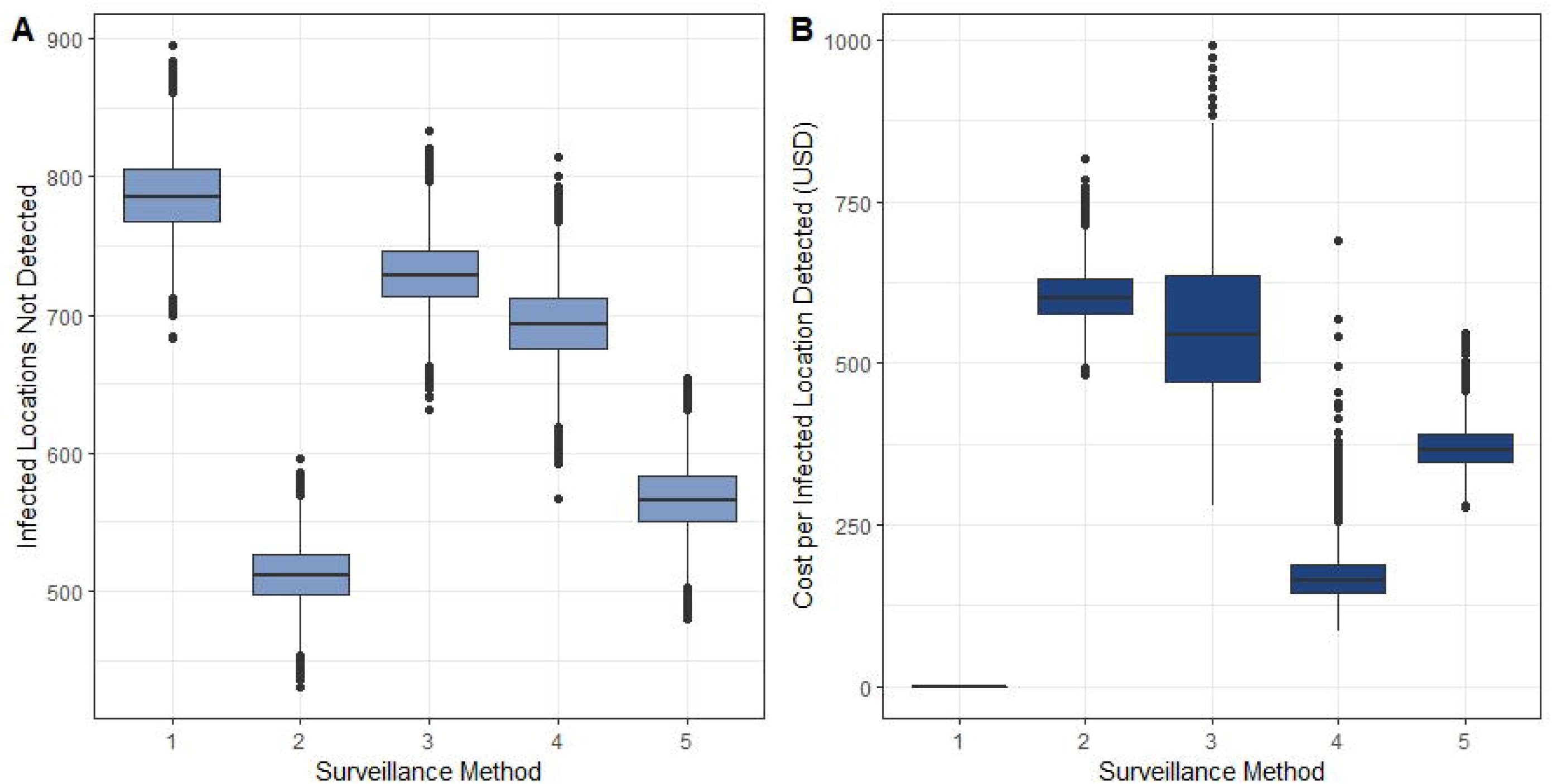
Evaluation of surveillance methods described in Table 3 by A. estimated numbers of locations with malaria infections not detected, and B. estimated additional costs per location with malaria infections detected (relative to standard PCD)

Based on distributions of malaria infections and probability of detection by routine diagnostics, we additionally explored the use of environmentally-stratified risk-based surveillance approaches. We defined high-risk areas based on distances from closed canopy forest (Figure S3). Evaluating costs of each surveillance system relative to baseline costs of standard PCD (Methods, SI), we estimated the total cost per location with malaria infection identified (Figure 3B). As the cost per infection detected was most sensitive to inclusion of molecular diagnostics, we defined a risk-based surveillance approach using health facility surveys in all areas and only applying molecular diagnostics to locations within 100m of closed canopy forest areas (Table 3, Figure S5). This risk-based surveillance approach almost halved the cost of detecting a location of infection compared to enhanced surveillance, from USD 603.10 (95% BCI: 530.02 - 690.82) to USD 370.00 (95% BCI: 313.18- 444.04) while detecting almost as many locations of infections.

**Table 3.** Surveillance methods assessed

|  | Survey method |  | Diagnostic method |  | Total cost (USD) |
| --- | --- | --- | --- | --- | --- |
|  | Passive case detection | Health facility surveys | Routine (RDT/ microscopy) | Molecular |  |
| 1: Standard PCD | X |  | X |  | - |
| 2: Enhanced surveillance | X | X | X | X | 193,547.70 |
| 3: PCD + molecular | X |  | X | X | 56,654.40 |
| 4: Health facility surveys + routine | X | X | X |  | 22,844.50 |
| 5: Risk-based surveys + diagnostics | X | Risk zone only | X | Risk zone only |  |

## Discussion

The spatial distribution of malaria is driven by a complex interplay of environmental and social factors influencing both disease transmission and identification by health systems. Statistical methods enabling examination of processes driving infection and detection provide an invaluable tool to evaluate the roles of environmental factors and develop targeted surveillance approaches. Here, we demonstrate a convenience sampling approach using health facility surveys markedly increases spatial coverage of surveillance systems; incorporating these surveys with satellite-derived remote sensing data allows estimation of the underlying distribution of infection not captured by passive surveillance. We additionally show higher proportions of subpatent malaria infections in forested areas, highlighting the limited utility of routine diagnostics within these regions. Applying these findings, we develop a cost-effective and operationally feasible risk-based surveillance approach using environmental data and illustrate how landscape data can be incorporated into disease surveillance systems.

Despite extensive research identifying proximity to forests as risk factors for malaria infection in Southeast Asia (e.g. (3, 22-27)), landscape data are not routinely used to design or inform surveillance systems. Malaria control programmes typically conduct community-based active case detection in response to reported malaria cases (2); however, we show this may miss a substantial proportion of active malaria foci due to biases in health-seeking behaviour and increased prevalence of subpatent malaria within higher transmission areas. Although mechanisms driving this relationship between forest cover and subpatent malaria are not known, patent malaria infections are more common in children in this area and settlements in closer proximity to forests may have different demographic compositions (e.g. logging and plantation camps) (21). Previous studies have also suggested a role for immunity in high transmission areas, with individuals repeatedly exposed to malaria commonly having lower parasite densities (4). Despite lower parasite densities, these subpatent infections can lead to infections in mosquitoes and may have a critical role in sustaining transmission in elimination settings, highlighting the importance of identifying and treating these individuals (28).

While screening entire populations may be prohibitively costly and intensive, the increasing availability of satellite-based remote sensing data provides new opportunities to use environmental data to target surveillance activities (29). Surveillance systems for malaria, as well as for other low incidence or emerging diseases, are challenged by the need to identify relatively rare events with shifting spatial patterns. Widely used in veterinary epidemiology, risk-based surveillance uses known risk factors to focus intensive surveillance activities on the populations where rare events are most likely to occur (30). Integrating remote-sensing derived environmental data into this approach provides an adaptable framework which can be easily updated in changing landscapes. Further, quantifying the detection probabilities associated with these surveillance approaches allows estimation of biases in data and identification of populations not captured by opportunistic sampling and can be used to plan active case detection. For this example, using health facility surveys, this enables evaluation of spatial patterns of health seeking behaviour as well as populations outside health system coverage. Screening all individuals attending health facilities, including those accompanying patients or for nonfebrile illnesses, vastly increases the spatial coverage of surveillance. The improved performance of models including a common spatial term for both infection and detection suggests processes determining healthcare coverage also influence infection risks, demonstrating the multiple utilities of measuring health seeking behaviour in this context.

Within this study, remote sensing data was used not only as covariates but also to define the populations at risk. The application of machine learning approaches to identify building footprints from very high resolution satellite imagery has produced increasingly accurate estimates of household distributions and allows estimation of spatial distribution of the populations (31). Development of tablet-based methods including population data and satellite imagery enabled near real-time identification of the residences of health facility attendees in a rural population with no formal addresses and limited internet connectivity (11). These tools and datasets can be further expanded to create accessible interfaces for local health workers to use environmental and spatial data and incorporate risk-based decision pathways on screening procedures and diagnostic tests based on household locations and travel history. This approach can also be easily modified to include multiple diseases with different underlying environmental risk factors and updated to include new environmental data, such as near real time deforestation alerts (32). Despite advances in using meteorological data to forecast vector-borne disease epidemics, landscape data is rarely used operationally and may provide more actionable information within rapidly changing environments. While we have focused on malaria, this is additionally relevant for targeting risk-based surveillance of emerging diseases, such as by detecting rare spillover events in changing habitats.

Despite the utility of these methods, there were several limitations to this study. As this study was designed to identify spatial locations of malaria infections within the sampling year, we did not explore temporal patterns of infection or health seeking behaviours. However, the modelling approach used is easily extendable to incorporate dynamic state-space models of changes in infection over time (12). If health facility surveys were collected over longer periods of time, this could additionally be expanded to incorporate seasonally varying meteorological data to further refine risk stratifications. While populations at risk were defined using multiple datasets, this is likely to have limited coverage of highly mobile indigenous populations not residing in permanent structures. Future work could explore the utility of satellite imagery to identify these populations, such as through monitoring of forest disturbance or modelling movement patterns. Within this study, the numbers of infections with different *Plasmodium* species precluded species-specific analyses. As all species are transmitted by the same mosquito vectors, environmental risk factors are expected to be similar; however, future work could examine differences between spatial distribution by malaria species. Additionally, while molecular approaches for malaria are not easily applied in rural settings, new diagnostics, such as lateral flow assays and serological tests, may facilitate point of contact testing in the future (33, 34).

Even with these limitations, this provides a novel and adaptable surveillance approach for environmentally driven diseases and demonstrates the role of landscapes in driving malaria infection and detection within this region. Incorporation of forest data enables identification of cost-effective risk-based surveillance approaches which increase probabilities of detecting malaria infections and can be applied to support elimination efforts. Additionally, the process-based ecological modelling method used provides a flexible framework to quantify detection probabilities and estimate the true spatial distribution of infection using biased data from convenience-based sampling approaches.

## Methods

### Survey approaches

We conducted cross-sectional health facility-based surveys at 27 facilities, including at one Regional Health Unit (RHU), 9 Barangay Health Stations (BHS) and 17 RDT centres based in community health worker households, established to increase access to malaria diagnosis and treatment (Figure S1). For one week every month between June 2016 - June 2017, all individuals attending the health facility, regardless of symptoms or patient status, were asked to participate in this survey. For consenting individuals, malaria status was assessed using either microscopy or rapid diagnostic tests (RDTs), with finger-prick blood samples collected on Whatman 3MM filter paper for subsequent polymerase chain reaction (PCR) (21). We classified malaria as positive for any *Plasmodium* species as 75% of identified species were *P. falciparum* and all malaria species are transmitted by common vectors in this area. Household locations were identified on offline maps and basic demographic information was recorded using GeoODK on Android tablets (11).

### Spatial and environmental covariates

To define the locations of all households within Rizal, we extracted information on household structure locations from the Facebook High Resolution Settlement Layer, a 30m resolution satellite-based remote sensing derived dataset on all inhabited structures (35). This dataset was combined with all reported households identified by survey participants and geolocated households from the 2015 Philippine census (36). We resampled all datasets to 50m resolution and removed duplicate locations to account for spatial resolution of datasets and inaccuracies of reported household locations. To estimate detection probabilities, we classified households included by a surveillance scenario if one or more individuals were sampled. Similarly, households were classified as infected if one or more individuals were identified as infected by diagnostics used by the surveillance approach.

Plausible covariates used to model detection or infection probabilities were assembled (Table SI). Handheld GPS devices (Garmin, USA) were used to record locations of all sampled clinics and roads within the region. Travel time to the nearest sampled clinic was calculated as accumulated cost from friction surfaces extracted from (37). Additional environmental and spatial covariates included population density (38), Euclidean distance from roads and bioclimatic variables (39). Elevation and topographic measures, including topographic wetness index (TWI), upslope area and aspect, were calculated from the ASTER Global Digital Elevation Model (40). Forest cover was classified as over 50% canopy cover based on (18) and Euclidean distance was calculated to the forest edge, recent deforestation within the past year and historical deforestation within the previous five years. We additionally included closed canopy forest, defined as canopy cover over 90% with a minimum area of 0.5ha (41).

### Occupancy modelling

Covariates were extracted for all identified household locations and Pearson correlation analysis was used to assess multicollinearity between variables. To select variables for inclusion, nonspatial binomial generalised linear models were fit separately for detection probabilities and infection probabilities for each surveillance scenario using backwards stepwise model selection with a five-point threshold for improvement in deviance information criteria (DIC) to minimise overfitting (42). Residual spatial autocorrelation was assessed using Moran’s I and performance assessed by area under the receiver operating curve (AUC). Weakly informative priors of Normal (0, 1/0.01) were used for all intercepts and coefficients. All models were implemented in Integrated Nested Laplace (INLA), with 10,000 posterior samples generated from the approximated posterior distribution to include uncertainty in these estimates (43). Final models had an AUC of 85% for both surveillance approaches.

We modelled the distribution of infections under each surveillance scenario separately using occupancy models in which the probability of detecting an infection (*y_i_*) in location *i* is dependent on the probability of detection (*p_i_*) and presence of infection (*ω_i_*) (13), modelled as:

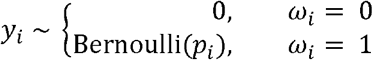

Where the linear predictor determining the probability of detection is modelled as:

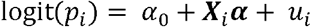

Where *α*_0_ represents the intercept, ***X_t_****α* represents a vector of covariate effects and *u_i_* is the spatial effect modelled as a Matern covariance function using the stochastic partial differential equations approach to represent the spatial process by Gaussian Markov random fields as implemented in INLA (43, 44). The process determining the true state of malaria presence ω is determined by the true probability of infection *ψ*:

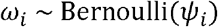

With the linear predictor for the Bernoulli model specified as:

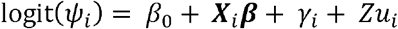

Where *β*_0_ represents the intercept, ***X_i_β*** represents a vector of covariate effects and *γ_i_* represents the spatial effect, modelled as described above. As processes influencing probability of detection (healthcare access) additionally may impact infection, we include a shared spatial component *Zu_i_* with scaling parameter *Z* (45).

### Patent malaria distribution

To explore factors affecting the distribution of locations of patent malaria infections (detected by RDT or microscopy) compared to all infections, we subset all malaria infected locations. For *J_j_* malaria infected individuals identified in each location, the number of patent infections observed (*m_i_*) is modelled as:

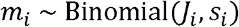

With the linear predictor determining the probability of patent infections (*s_i_*) modelled as:

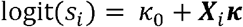

Where *κ*_0_ represents the intercept and ***X_i_κ*** represents a vector of covariate effects. Using data from all locations included in the study site, we then predicted a location specific sensitivity of routine diagnostics. Based on these results, we used environmental data to define an area with higher probabilities of malaria infections only detectable by molecular diagnostics.

### Evaluation of surveillance systems

We modelled the true probability of infection from the infection process model using data from all available diagnostics. To compare surveillance methods with different survey and diagnostic methods, we estimated the number of infected locations not detected as:

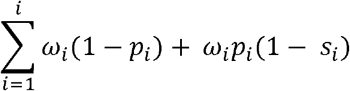

Where *p_i_*, is the probability of detection using different survey methods and *s_i_*, represents diagnostic sensitivity, with PCR considered the gold standard. We additionally included risk-based surveillance methods, using health facility surveys and molecular diagnostics in high risk areas defined by proximity to closed canopy forest. All derived quantities were estimated using 10,000 posterior samples.

To evaluate the cost effectiveness of different surveillance approaches, we estimated the additional costs to health systems of including different survey and diagnostic approaches (Table S4). This excluded capital costs and costs already covered by existing health systems (e.g. RDT and microscopy diagnostics for febrile patients). Health facility survey costs included additional payments to personnel, training, equipment, RDT and microscopy for non-febrile participants, sample collection and molecular diagnostics for all attendees and salary for a technician to support data management and sample analysis. The costs of molecular diagnostics included DNA extraction and PCR reagents, assuming all DNA extraction was completed using Chelex with 10% of samples verified using a commercial Qiagen kit. To account for varying numbers of samples screened from each location, we estimated the mean cost of molecular diagnostics per location as the total cost of molecular diagnostics divided by the total number of locations included. We evaluated these against the estimated number of locations correctly identified as a measure of cost effectiveness. All analysis was completed in R statistical programming language (v 3.6), with maps visualised in R or ArcGIS (ESRI, Redlands, USA).

### Ethics approval

This study was approached by the Institutional Review Board of the Research Institute for Tropical Medicine, Department of Health Philippines (IRB:2016-04) and the Research Ethics Committee of the London School of Hygiene and Tropical Medicine (11597). Written informed consent was obtained from all participants or parents or guardians and assent obtained from children under 18.

### Data availability

Data is available upon reasonable request and with permission of ethics committees in the Philippines and United Kingdom. All R code needed to conduct these analyses will be available at https://github.com/kfornace

## Data Availability

The datasets analysed and generated during this study are not publicly available due to ethical restrictions on sharing identifiable health data. Data are available from authors upon reasonable request and with permission of relevant institutional review boards.

## Acknowledgements

We would like to acknowledge the Newton Fund, Philippine Council for Health Research and Development and UK Medical Research Council for funding received for ENSURE: Enhanced Surveillance for control and elimination of malaria in the Philippines. We would additionally like to thank Ellaine de la Fuente and Beulah Boncayo for supporting fieldwork as well as the local health staff and government of Rizal, Palawan, the Philippines who supported this survey.

## Author contributions

KMF, FE, JRCH and CJD designed this study. KMF analysed the data and wrote the manuscript. KMF and RAR collected and analysed spatial and remote sensing data. RAR, MLMM, APNB and JSL collected the health data and analysed samples. All authors read and approved the final manuscript.

